# Correlates of COVID-19 conspiracy theory beliefs in Japan: A nationally-representative cross-sectional survey

**DOI:** 10.1101/2024.09.06.24313228

**Authors:** Yukihiro Sato, Ichiro Kawachi, Yasuaki Saijo, Eiji Yoshioka, Ken Osaka, Takahiro Tabuchi

## Abstract

**Background:** The COVID-19 pandemic was associated with an increase in conspiracy theories worldwide. However, the prevalence of COVID-19 conspiracy beliefs among Japanese has remained unclear. This study aimed to estimate the prevalence and correlates of COVID-19 conspiracy beliefs in Japan using a nationwide survey of 28,175 residents aged 16–81 years old.

**Methods:** A nationally representative, cross-sectional self-administered survey was conducted from September to October 2021. To assess the number of COVID-19 conspiracy beliefs, we used three questions from the Oxford Coronavirus Explanations, Attitudes, and Narratives Survey. Independent variables included general vaccine conspiracy beliefs, sociodemographic variables, information sources for COVID-19, trust in authorities, and fear of COVID-19.

**Results:** After applying sampling weights and imputation, the estimated prevalence of holding at least one COVID-19 conspiracy belief was 24.4%. From a linear regression model, several factors were independently associated with conspiracy beliefs. Notably, people with the lowest level of education (lower secondary school) endorsed fewer COVID-19 conspiracy beliefs (B -0.089, vs. upper secondary school). Furthermore, higher socioeconomic backgrounds–such as higher income, higher wealth, and regular employment–were associated with endorsing conspiracy beliefs. Additionally, only 37.3% of respondents trusted the government of Japan, but paradoxically, trust in the government was positively associated with conspiracy beliefs (B 0.175, vs. distrust).

**Conclusions:** Conspiracy beliefs about COVID-19 were prevalent in about a quarter of the Japanese population. Certain groups are more likely to endorse conspiracy beliefs, and targeting interventions towards these groups might be efficient in stemming the spread of conspiracy beliefs.

## INTRODUCTION

A simple definition of conspiracy theory is “an explanation that a significant event has resulted from a small group of powerful actors engaging in malevolent activities, which is not officially recognised as a salient cause” [1–5]. Beliefs in conspiracy theories have proliferated; especially, social crises stimulate and increase the prevalence [6,7]. Since late December 2019, misinformation and conspiracy theories about the coronavirus disease (COVID-19) pandemic have been spreading [6,8]. For example, there are various conspiracy theories about COVID-19: the virus was created to reduce the global population; pharmaceutical companies created and spread the virus to gain profit [9]. The worldwide prevalence of different types of conspiracy beliefs about COVID-19 varied from 5.0% to 39.0% [9,10]. Most of the studies were from the US and European countries, but there were no studies on the prevalence of conspiracy beliefs about COVID-19 in Japan. Conspiracy theories about COVID-19 have undermined public health measures and induced a threat to people’s health. Previous studies indicated that people who believe in conspiracy theories tended to be reluctant to get vaccinated and to disregard guidelines, such as social distancing, hand washing, and wearing a face mask [10–14]. The World Health Organization (WHO) has called for active steps to combat the “infodemic” [6,8].

Accumulated evidence has shed light on predictors of conspiracy beliefs about COVID-19 [10,12,14–16]. Prior research has indicated that belief in one conspiracy theory predicts holding other conspiracy beliefs simultaneously even if they are contradictory [17–20]. The reasons are considered to be a conspiratorial mindset, which is defined as a tendency to favour conspiracy explanations due to a bias against powerful actors and authorities [17–20]. Demographic variables, such as age and sex, are associated with the endorsement of COVID-19 conspiracy beliefs, but prior studies show conflicting results [10,14]. The variation might be due to cultural backgrounds [14]. Socioeconomic status, such as income and education levels, predict conspiracy beliefs well [10,14]. In particular, individuals with lower educational attainment are more likely to believe the COVID-19 conspiracy theory [21]. The associations might be mediated through the interplay of thinking style and feelings of control [21]. Furthermore, exposure to low-level information might also be one of the explanation factors [14]. Conspiracy beliefs regarding COVID-19 are abundant on the internet, especially on social media. People shared conspiracy theories on a variety of media forms, such as YouTube, Instagram, and Twitter (currently known as X), and conspiracy theories were reinforced and amplified [6,15]. Social media is considered one source of conspiracy theories going viral [10,14,15], including Japan [22]. Furthermore, distrust in authorities hampers scientific communication. Scientific information is not directly transmitted from scientists, but relayed by governments and national health institutions. Trust in authorities can play an important role [10,14]. Feelings of fear and anxiety were associated with beliefs in conspiracy theories [10,14], because conspiracy theories can be interpreted as expressions of people’s fear and anxieties [16]. Further, experiencing discrimination based on infections of COVID-19 can facilitate endorsing conspiracy beliefs. Indeed, in cases of HIV, British White gay men who experienced discrimination tended to hold HIV conspiracy theories [12,23].

As in other countries, COVID-19 conspiracy theories have proliferated in Japan. A Trump-inspired party (Sansei to), which advanced conspiracy theories about COVID-19 (e.g. laboratory origin narrative and big Pharma plot narratives), secured a seat in the House of Councillors in 2022 [24]. Conspiracy theories appear to be gaining popularity in Japan. However, the prevalence of conspiracy theories about COVID-19 has remained unclear. In addition, Japan has a different situation from other countries. Previous studies have found that individuals who rely on traditional media tend to be less inclined to believe in conspiracy theories [25,26]. However, even in traditional media such as TV news and newspapers, there is a history of biased health reporting in Japan. For example, Japan’s human papillomavirus vaccination rate is low, due to excessive reports of false side effects by traditional media [27]. This situation might have facilitated distrust in government officials, healthcare professionals, and scientists. Indeed, Japan had relatively low trust in the government in 2020 among OECD countries [28]. However, there is a scarcity of research conducted in Japan focusing on correlates of conspiracy beliefs in COVID-19. This study aimed to estimate the prevalence of conspiracy beliefs about COVID-19 and explore factors associated with holding conspiracy beliefs about COVID-19 among a Japanese nationally representative sample.

## METHODS

### Ethics approval and consent to participate

The study protocol was reviewed and approved by the Research Ethics Committee of the Osaka International Cancer Institute (approved on June 19, 2020; approval number 20084). The authors assert that all procedures contributing to this work comply with the ethical standards of the relevant national and institutional committees on human experimentation and with the Helsinki Declaration of 1975, as revised in 2008. All participants provided written informed consent online before responding to the questionnaire. Participants received Rakuten points as a reward, which can be used for online shopping. The exact number of points is not disclosed at the request of an internet research agency (Rakuten Insight, Inc., Tokyo, Japan).

### Study design, data sources, and participants

This cross-sectional study used a dataset from the Japan COVID-19 and Society Internet Survey (JACSIS) (https://jacsis-study.jp/), which is a nationally representative web-based self-administered questionnaire survey. The JACSIS conducts a survey annually and targets approximately 2 million panellists, registering with an internet research agency (Rakuten Insight, Inc., Tokyo, Japan). In the JACSIS, the target population was individuals aged 15–79 years who lived in Japan at a survey. The survey in this study was launched on 27 September 2021 and concluded on 29 October 2021. Of all previous respondents of the previous wave of the JACSIS, an Internet research agency followed up with 33,081 individuals who were available to respond. From 27 September to 29 October 2021, a follow-up survey was conducted among them, resulting in responses from 22,838 individuals (response rate = 69%). Additionally, to achieve a total of 31,000 participants, an additional survey using the same questionnaire was conducted from 23 to 28 October 2021. Consequently, an extra 8,162 responses were collected.

Participants who provided fraudulent responses were excluded to ensure the quality of the data. Three exclusion criteria were employed to identify fraudulent responses: an incorrect answer to an attention check question (“Please select the second last option from the following choices.”), straight-line responses regarding drug use, and regarding comorbidities, leading to the exclusion of 2,825 respondents. Thus, the Analytic population was 28,175.

### Dependent variable: The number of conspiracy beliefs regarding COVID-19

To assess conspiracy beliefs about COVID-19, we used three questions from the Oxford Coronavirus Explanations, Attitudes, and Narratives Survey (OCEANS) [29]. The questions were as follows: big Pharma created COVID-19 to profit from the vaccines; COVID-19 was created to force everyone to get vaccinated; the vaccine will be used to carry out mass sterilisation. The available response options were “1. Strongly agree”, “2. Somewhat agree ”, “3. Neither agree nor disagree”, “4. Somewhat disagree”, and “5. Strongly disagree”. We defined the first and second options as believing in conspiracy theories about COVID-19 vaccines. The Cronbach’s alpha was 0.889 and the Pearson’s correlation matrix of the measure is shown in Appendix Table 1.

### Independent variables

Based on previous studies [10,12,14–16], we selected variables potentially associated with COVID-19 conspiracy beliefs: general vaccine conspiracy beliefs, sociodemographic variables, information sources for COVID-19, trust in authorities, fear of COVID-19, and others.

#### 1. General vaccine conspiracy beliefs

We calculated the number of conspiracy beliefs about general vaccines using a Vaccine Conspiracy Beliefs Scale (VCBS) involving 7 items [30]. The questions of VCBS were below: vaccine safety data is often fabricated; immunizing children is harmful and this fact is covered up; pharmaceutical companies cover up the dangers of vaccines; people are deceived about vaccine efficacy; vaccine efficacy data is often fabricated; people are deceived about vaccine safety; the government is trying to cover up the link between vaccines and autism. Potential responses were a 7-point scale that ranged from “1. strongly disagree” to “7. strongly agree”. We defined the fifth to seventh options as believing conspiracy theories about general vaccines. The Cronbach’s alpha was 0.947 and the Pearson’s correlation matrix of the measure is shown in Appendix Table 2.

#### 2. Sociodemographic variables

We included age (16–19, 20–24, 25–29, 30–34, 35–39, 40–44, 45–49, 50–54, 55–59, 60–64, 65–69, 70–74, and 75–81 years), sex (men and women), marital status (married, unmarried, widowed, and divorced), educational attainment (lower secondary school, upper secondary school, specialised training college [post-secondary courses], junior college and college of technology, university, and master’s or doctor’s degree), employment status (regular employee, temporary employee, self-employed, employer, student, unemployed or retired, housekeeper, and others), annual household income (0 to <2, 2 to <4, 4 to <5, 5 to <8, and ≥8 million yen), household financial assets (0 to <1, 1 to <4, 4 to <9, 9 to <20, and ≥20 million yen), and household indebtedness (none, >0 to <2, and ≥2 million yen).

#### 3. Information sources for COVID-19

As for sources of information about COVID-19, we targeted the following 14 information sources: websites of government agencies, websites of research institutions, video sharing platforms (e.g. YouTube), LINE, Twitter, Facebook, Instagram, web news, newspapers, magazines, books, TV news, and tabloid TV shows. Sources of information for COVID-19 were assessed by the following question: “Have you obtained medical information about health topics, including COVID-19, from each of the following sources?” The options available for each source were “Yes” and “No”. If respondents chose “Yes”, they were asked that “How much do you trust the source of information you selected in the previous question?” The possible options were “1. Very trust”, “ 2. Trust”, “3. Somewhat trust”, “4. Somewhat distrust”, ”5. Distrust”, and “ 6. Distrust at all”. We defined the first to third options as “trusted”, and the fourth to sixth as “distrusted”. Combining the former and latter questions, we defined three categories: “Not use”, “Use but distrust”, and “Use and trust”.

#### 4. Trust in authorities

To measure the level of trust in authorities, we targeted the following 3 authorities: the government of Japan, prefectural administrations, and municipal administrations. Trust in authorities was measured using the following questions: the government is trustworthy; the administration of the municipality in which you currently live is trustworthy; the administration of the prefecture in which you currently live is trustworthy. The response options were “1. Agree”, “2. Somewhat agree”, “3. Somewhat disagree”, and “4. Disagree”. We defined the first and second options as “Trust”, and the third and fourth as “Distrust”.

#### 5. Fear of COVID-19

We used the Japanese Version of the Fear of Coronavirus Disease 2019 Scale (FCV-19S) [31]. This scale involves 7 items with a scale of 1 (strongly disagree) to 5 (strongly agree) points. The range of the scale was from 7 to 35 points. We adopted ≥21 of a cut-off point to identify psychological distress or difficulties in daily living due to the fear of COVID-19 [32]. The Cronbach’s alpha was 0.836 and the Pearson’s correlation matrix of the measure is shown in Appendix Table 3.

#### 6. Other covariates

Experiencing discrimination regarding COVID-19 was obtained using a question: “In the last two months, have you experienced any of the following events? Felt discrimination related to COVID-19.” Available options were “1. Yes (experienced for the first time in the last two months)”, “2. Yes (and had happened before)”, “3. Never (but had happened before)”, and “4. Never happened before”. We defined the first to third options as “Experienced”, and the last option as “Never”. Additionally, we obtained a medical history of depression, other mental disorders, and coronavirus infections.

### Statistical analysis

We employed linear regression analysis with a robust error variance to estimate the unstandardised coefficients (Bs) of the number of conspiracy beliefs regarding COVID-19. Although the distribution of a dependent variable is right-skewed, linear regression models can provide valid estimations [33]. A positive coefficient means holding more conspiracy theories while a negative coefficient means holding fewer conspiracy theories. We created two models: a crude model and a fully adjusted model using the simultaneous forced-entry method. The fully adjusted model included all independent variables simultaneously. To confirm the validity of the results from linear regression models, we also conducted a Poisson regression analysis with a robust error variance to estimate prevalence ratios for holding at least one conspiracy belief in COVID-19 [34].

To impute missing values, we used a k-nearest neighbour imputation (the R package “VIM”) [35]. After the imputation, sampling weights were calculated using the raking method from the R package “anesrake” to match the proportions of age, sex, prefecture of residence, marital status, annual household income, and educational attainment from census data. Detailed information on the sampling weights has been described in Appendix Table 4. A P-value <0.05 (2-tailed) was considered statistically significant. All analyses were conducted in R (ver. 4.3.0; R Foundation for Statistical Computing, Vienna, Austria).

## RESULTS

Table 1 shows basic participant characteristics. The median age of respondents was 50, ranging from 16 to 81. The proportion of women was 50.8%. After imputation and weighting, 10.6% (95% confidence interval [CI] 10.1, 11.1) of the participants believed in one conspiracy theory regarding COVID-19 (OCEANS), 6.5% (95%CI 6.1, 6.9) in two, and 7.3% (95%CI 7.0, 7.8) in three. The prevalence of holding at least one conspiracy theory about COVID-19 was 24.4% (95%CI 23.7, 25.1). The detailed results of the original categories for three questions on COVID-19 conspiracy beliefs are shown in Appendix Table 5. On the other hand, participants who believed in at least one conspiracy theory regarding general vaccines (VCBS) constituted 33.6% (detailed results are shown in Appendix Table 6). The main source of COVID-19 information was TV news, trusted by 67.9% of the participants. Web news, the second source, garnered 48.0% trust. Distrust in the Japanese government was expressed by 62.7% of the participants.

**Table 1.** Basic characteristics of 28,175 participants.

| Variable | Response participants |  | Imputed population |  | Weighted imputed population |  |
| --- | --- | --- | --- | --- | --- | --- |
|  | n=28,175 | (%) | n=28,175 | (%) | n=28,175 | (%) |
| **Dependent variable** |  |  |  |  |  |  |
| The number of conspiracy theory beliefs regarding COVID-19 |  |  |  |  |  |  |
| 0 | 21,414 | (76.0%) | 21,414 | (76.0%) | 21,297 | (75.6%) |
| 1 | 2,876 | (10.2%) | 2,876 | (10.2%) | 2,974 | (10.6%) |
| 2 | 1,830 | (6.5%) | 1,830 | (6.5%) | 1,836 | (6.5%) |
| 3 | 2,055 | (7.3%) | 2,055 | (7.3%) | 2,068 | (7.3%) |
| **Independent variable** |  |  |  |  |  |  |
| The number of conspiracy theory beliefs regarding general vaccines |  |  |  |  |  |  |
| 0 | 18,798 | (66.7%) | 18,798 | (66.7%) | 18,712 | (66.4%) |
| 1 | 3,021 | (10.7%) | 3,021 | (10.7%) | 3,010 | (10.7%) |
| 2 | 2,022 | (7.2%) | 2,022 | (7.2%) | 2,067 | (7.3%) |
| 3 | 1,180 | (4.2%) | 1,180 | (4.2%) | 1,203 | (4.3%) |
| 4 | 873 | (3.1%) | 873 | (3.1%) | 865 | (3.1%) |
| 5 | 772 | (2.7%) | 772 | (2.7%) | 768 | (2.7%) |
| 6 | 741 | (2.6%) | 741 | (2.6%) | 779 | (2.8%) |
| 7 | 768 | (2.7%) | 768 | (2.7%) | 770 | (2.7%) |
| Age (years old) |  |  |  |  |  |  |
| 16–19 | 573 | (2.0%) | 573 | (2.0%) | 949 | (3.4%) |
| 20–24 | 1,829 | (6.5%) | 1,829 | (6.5%) | 1,801 | (6.4%) |
| 25–29 | 1,797 | (6.4%) | 1,797 | (6.4%) | 1,834 | (6.5%) |
| 30–34 | 1,963 | (7.0%) | 1,963 | (7.0%) | 1,885 | (6.7%) |
| 35–39 | 2,186 | (7.8%) | 2,186 | (7.8%) | 2,114 | (7.5%) |
| 40–44 | 2,600 | (9.2%) | 2,600 | (9.2%) | 2,350 | (8.3%) |
| 45–49 | 2,854 | (10.1%) | 2,854 | (10.1%) | 2,798 | (9.9%) |
| 50–54 | 2,529 | (9.0%) | 2,529 | (9.0%) | 2,661 | (9.4%) |
| 55–59 | 2,256 | (8.0%) | 2,256 | (8.0%) | 2,250 | (8.0%) |
| 60–64 | 2,234 | (7.9%) | 2,234 | (7.9%) | 2,125 | (7.5%) |
| 65–69 | 2,648 | (9.4%) | 2,648 | (9.4%) | 2,263 | (8.0%) |
| 70–74 | 2,655 | (9.4%) | 2,655 | (9.4%) | 2,781 | (9.9%) |
| 75–81 | 2,051 | (7.3%) | 2,051 | (7.3%) | 2,362 | (8.4%) |
| Sex |  |  |  |  |  |  |
| Men | 13,870 | (49.2%) | 13,870 | (49.2%) | 13,986 | (49.6%) |
| Women | 14,305 | (50.8%) | 14,305 | (50.8%) | 14,189 | (50.4%) |
| Marital status |  |  |  |  |  |  |
| Married | 16,969 | (60.2%) | 16,969 | (60.2%) | 17,472 | (62.0%) |
| Unmarried | 8,322 | (29.5%) | 8,322 | (29.5%) | 7,918 | (28.1%) |
| Widowed | 1,001 | (3.6%) | 1,001 | (3.6%) | 1,200 | (4.3%) |
| Divorced | 1,883 | (6.7%) | 1,883 | (6.7%) | 1,585 | (5.6%) |
| Educational attainment |  |  |  |  |  |  |
| Lower Secondary School | 392 | (1.4%) | 392 | (1.4%) | 1,960 | (7.0%) |
| Upper Secondary School | 7,592 | (27.1%) | 7,638 | (27.1%) | 12,780 | (45.4%) |
| Specialised Training College (Post- | 3,203 | (11.4%) | 3,230 | (11.5%) | 2,775 | (9.9%) |
|  | n=28,175 | (%) | n=28,175 | (%) | n=28,175 | (%) |
| *Secondary Courses)* |  |  |  |  |  |  |
| *Junior College and College of Technology* | 3,045 | (10.9%) | 3,065 | (10.9%) | 2,447 | (8.7%) |
| *University* | 12,368 | (44.2%) | 12,440 | (44.2%) | 7,421 | (26.3%) |
| *Master's or doctor's degree* | 1,409 | (5.0%) | 1,410 | (5.0%) | 791 | (2.8%) |
| *(Missing)* | 166 |  |  |  |  |  |
| **Employment status** |  |  |  |  |  |  |
| *Regular employee* | 9,722 | (34.5%) | 9,722 | (34.5%) | 9,231 | (32.8%) |
| *Temporary employee* | 4,881 | (17.3%) | 4,881 | (17.3%) | 4,976 | (17.7%) |
| *Self-employed* | 1,732 | (6.1%) | 1,732 | (6.1%) | 1,659 | (5.9%) |
| *Employer* | 990 | (3.5%) | 990 | (3.5%) | 980 | (3.5%) |
| *Student* | 1,267 | (4.5%) | 1,267 | (4.5%) | 1,387 | (4.9%) |
| *Unemployed or retired* | 4,636 | (16.5%) | 4,636 | (16.5%) | 4,768 | (16.9%) |
| *Housekeeper* | 4,711 | (16.7%) | 4,711 | (16.7%) | 4,938 | (17.5%) |
| *Others* | 236 | (0.8%) | 236 | (0.8%) | 235 | (0.8%) |
| **Annual household income** |  |  |  |  |  |  |
| *0 to <2 million yen* | 2,415 | (10.9%) | 2,575 | (9.1%) | 3,298 | (11.7%) |
| *2 to <4 million yen* | 5,738 | (25.9%) | 7,142 | (25.3%) | 8,298 | (29.5%) |
| *4 to <5 million yen* | 2,868 | (13.0%) | 3,908 | (13.9%) | 3,669 | (13.0%) |
| *5 to <8 million yen* | 6,082 | (27.5%) | 8,530 | (30.3%) | 7,219 | (25.6%) |
| *≥8 million yen* | 5,024 | (22.7%) | 6,020 | (21.4%) | 5,692 | (20.2%) |
| *(Missing)* | 6,048 |  |  |  |  |  |
| **Household financial assets** |  |  |  |  |  |  |
| *0 to <1 million yen* | 3,034 | (17.5%) | 4,330 | (15.4%) | 5,144 | (18.3%) |
| *1 to <4 million yen* | 3,779 | (21.8%) | 6,707 | (23.8%) | 7,434 | (26.4%) |
| *4 to <9 million yen* | 3,440 | (19.9%) | 6,594 | (23.4%) | 6,245 | (22.2%) |
| *9 to <20 million yen* | 3,247 | (18.7%) | 5,349 | (19.0%) | 4,931 | (17.5%) |
| *≥20 million yen* | 3,825 | (22.1%) | 5,195 | (18.4%) | 4,422 | (15.7%) |
| *(Missing)* | 10,850 |  |  |  |  |  |
| **Household indebtedness** |  |  |  |  |  |  |
| *None* | 15,722 | (71.5%) | 20,145 | (71.5%) | 19,914 | (70.7%) |
| *>0 to <2 million yen* | 1,510 | (6.9%) | 2,005 | (7.1%) | 2,236 | (7.9%) |
| *≥2 million yen* | 4,771 | (21.7%) | 6,025 | (21.4%) | 6,025 | (21.4%) |
| *(Missing)* | 6,172 |  |  |  |  |  |
| **Information source for COVID-19:** |  |  |  |  |  |  |
| **Websites of government agencies** |  |  |  |  |  |  |
| *Not use* | 15,467 | (54.9%) | 15,467 | (54.9%) | 16,182 | (57.4%) |
| *Use but distrust* | 827 | (2.9%) | 827 | (2.9%) | 850 | (3.0%) |
| *Use and trust* | 11,881 | (42.2%) | 11,881 | (42.2%) | 11,143 | (39.5%) |
| **Information source for COVID-19:** |  |  |  |  |  |  |
| **Websites of research institutions** |  |  |  |  |  |  |
| *Not use* | 25,179 | (89.4%) | 25,179 | (89.4%) | 25,468 | (90.4%) |
| *Use but distrust* | 217 | (0.8%) | 217 | (0.8%) | 224 | (0.8%) |
| *Use and trust* | 2,779 | (9.9%) | 2,779 | (9.9%) | 2,484 | (8.8%) |
| **Information source for COVID-19:** |  |  |  |  |  |  |
| **Video sharing platforms (e.g. YouTube)** |  |  |  |  |  |  |
|  | n=28,175 | (%) | n=28,175 | (%) | n=28,175 | (%) |
| *Not use* | 23,647 | (83.9%) | 23,647 | (83.9%) | 23,534 | (83.5%) |
| *Use but distrust* | 1,229 | (4.4%) | 1,229 | (4.4%) | 1,225 | (4.3%) |
| *Use and trust* | 3,299 | (11.7%) | 3,299 | (11.7%) | 3,416 | (12.1%) |
| Information source for COVID-19: LINE |  |  |  |  |  |  |
| *Not use* | 23,218 | (82.4%) | 23,218 | (82.4%) | 23,040 | (81.8%) |
| *Use but distrust* | 845 | (3.0%) | 845 | (3.0%) | 862 | (3.1%) |
| *Use and trust* | 4,112 | (14.6%) | 4,112 | (14.6%) | 4,273 | (15.2%) |
| Information source for COVID-19: Twitter |  |  |  |  |  |  |
| *Not use* | 24,019 | (85.2%) | 24,019 | (85.2%) | 24,080 | (85.5%) |
| *Use but distrust* | 1,431 | (5.1%) | 1,431 | (5.1%) | 1,348 | (4.8%) |
| *Use and trust* | 2,725 | (9.7%) | 2,725 | (9.7%) | 2,747 | (9.8%) |
| Information source for COVID-19: Facebook |  |  |  |  |  |  |
| *Not use* | 26,615 | (94.5%) | 26,615 | (94.5%) | 26,605 | (94.4%) |
| *Use but distrust* | 461 | (1.6%) | 461 | (1.6%) | 455 | (1.6%) |
| *Use and trust* | 1,099 | (3.9%) | 1,099 | (3.9%) | 1,115 | (4.0%) |
| Information source for COVID-19: Instagram |  |  |  |  |  |  |
| *Not use* | 26,474 | (94.0%) | 26,474 | (94.0%) | 26,362 | (93.6%) |
| *Use but distrust* | 493 | (1.7%) | 493 | (1.7%) | 518 | (1.8%) |
| *Use and trust* | 1,208 | (4.3%) | 1,208 | (4.3%) | 1,296 | (4.6%) |
| Information source for COVID-19: Web news |  |  |  |  |  |  |
| *Not use* | 9,828 | (34.9%) | 9,828 | (34.9%) | 10,350 | (36.7%) |
| *Use but distrust* | 4,556 | (16.2%) | 4,556 | (16.2%) | 4,308 | (15.3%) |
| *Use and trust* | 13,791 | (48.9%) | 13,791 | (48.9%) | 13,517 | (48.0%) |
| Information source for COVID-19: Newspapers |  |  |  |  |  |  |
| *Not use* | 15,986 | (56.7%) | 15,986 | (56.7%) | 16,996 | (60.3%) |
| *Use but distrust* | 829 | (2.9%) | 829 | (2.9%) | 772 | (2.7%) |
| *Use and trust* | 11,360 | (40.3%) | 11,360 | (40.3%) | 10,407 | (36.9%) |
| Information source for COVID-19: Magazines |  |  |  |  |  |  |
| *Not use* | 25,105 | (89.1%) | 25,105 | (89.1%) | 25,338 | (89.9%) |
| *Use but distrust* | 560 | (2.0%) | 560 | (2.0%) | 512 | (1.8%) |
| *Use and trust* | 2,510 | (8.9%) | 2,510 | (8.9%) | 2,325 | (8.3%) |
| Information source for COVID-19: Books |  |  |  |  |  |  |
| *Not use* | 25,840 | (91.7%) | 25,840 | (91.7%) | 26,140 | (92.8%) |
| *Use but distrust* | 267 | (0.9%) | 267 | (0.9%) | 254 | (0.9%) |
| *Use and trust* | 2,068 | (7.3%) | 2,068 | (7.3%) | 1,781 | (6.3%) |
| Information source for COVID-19: TV news |  |  |  |  |  |  |
| *Not use* | 5,770 | (20.5%) | 5,770 | (20.5%) | 6,261 | (22.2%) |
| *Use but distrust* | 2,892 | (10.3%) | 2,892 | (10.3%) | 2,778 | (9.9%) |
|  | n=28,175 | (%) | n=28,175 | (%) | n=28,175 | (%) |
| *Use and trust* | 19,513 | (69.3%) | 19,513 | (69.3%) | 19,136 | (67.9%) |
| Information source for COVID-19: |  |  |  |  |  |  |
| Tabloid TV shows |  |  |  |  |  |  |
| *Not use* | 11,502 | (40.8%) | 11,502 | (40.8%) | 11,572 | (41.1%) |
| *Use but distrust* | 3,710 | (13.2%) | 3,710 | (13.2%) | 3,517 | (12.5%) |
| *Use and trust* | 12,963 | (46.0%) | 12,963 | (46.0%) | 13,086 | (46.4%) |
| Trust in the government of Japan |  |  |  |  |  |  |
| *Distrust* | 17,596 | (62.5%) | 17,596 | (62.5%) | 17,674 | (62.7%) |
| *Trust* | 10,579 | (37.5%) | 10,579 | (37.5%) | 10,501 | (37.3%) |
| Trust in the prefectural administration |  |  |  |  |  |  |
| *Distrust* | 13,350 | (47.4%) | 13,350 | (47.4%) | 13,603 | (48.3%) |
| *Trust* | 14,825 | (52.6%) | 14,825 | (52.6%) | 14,572 | (51.7%) |
| Trust in the municipal administration |  |  |  |  |  |  |
| *Distrust* | 12,993 | (46.1%) | 12,993 | (46.1%) | 13,359 | (47.4%) |
| *Trust* | 15,182 | (53.9%) | 15,182 | (53.9%) | 14,816 | (52.6%) |
| Fear of COVID-19 |  |  |  |  |  |  |
| *None* | 18,059 | (64.1%) | 18,059 | (64.1%) | 17,626 | (62.6%) |
| *Feeling* | 10,116 | (35.9%) | 10,116 | (35.9%) | 10,549 | (37.4%) |
| Discriminated against related to COVID-19 |  |  |  |  |  |  |
| *Never* | 26,563 | (94.3%) | 26,563 | (94.3%) | 26,516 | (94.1%) |
| *Experienced* | 1,612 | (5.7%) | 1,612 | (5.7%) | 1,659 | (5.9%) |
| Medical history of COVID-19 |  |  |  |  |  |  |
| *None* | 27,663 | (98.2%) | 27,663 | (98.2%) | 27,568 | (97.8%) |
| *Diagnosed within the past year* | 260 | (0.9%) | 260 | (0.9%) | 348 | (1.2%) |
| *Diagnosed before the past year* | 252 | (0.9%) | 252 | (0.9%) | 258 | (0.9%) |
| Medical history of depression |  |  |  |  |  |  |
| *Never* | 24,691 | (87.6%) | 24,691 | (87.6%) | 24,603 | (87.3%) |
| *Former* | 2,136 | (7.6%) | 2,136 | (7.6%) | 2,101 | (7.5%) |
| *Current* | 1,348 | (4.8%) | 1,348 | (4.8%) | 1,470 | (5.2%) |
| Medical history of other mental disorders |  |  |  |  |  |  |
| *Never* | 25,647 | (91.0%) | 25,647 | (91.0%) | 25,527 | (90.6%) |
| *Former* | 1,229 | (4.4%) | 1,229 | (4.4%) | 1,317 | (4.7%) |
| *Current* | 1,299 | (4.6%) | 1,299 | (4.6%) | 1,331 | (4.7%) |

Table 2 presents the weighted mean and proportions of the number of conspiracy beliefs regarding COVID-19. Among age groups, participants aged 16–19 years had the highest mean of conspiracy beliefs regarding COVID-19 (the weighted mean was 0.56), followed by 20–24 years (0.53) and 75–81 years (0.51). Women had a lesser number of conspiracy beliefs than men (0.39 vs 0.52). Participants who used and trusted TV news had a smaller number of conspiracy beliefs (0.43) than ones who did not use (0.54) and distrusted (0.48). Participants who trusted web news had a smaller number of conspiracy beliefs (-0.44) than those who did not use it (0.50), but a larger number than those who distrusted it. Paradoxically, participants who distrusted the Japanese government endorsed fewer conspiracy beliefs (0.40) compared to those who trusted the government (0.56). By contrast, there was no difference in the number of conspiracy beliefs between participants who trusted versus distrusted their prefectural and municipal administrations, respectively (prefectural administration: trust 0.42, distrust 0.42; municipal administration: trust 0.42, distrust 0.42).

**Table 2.**
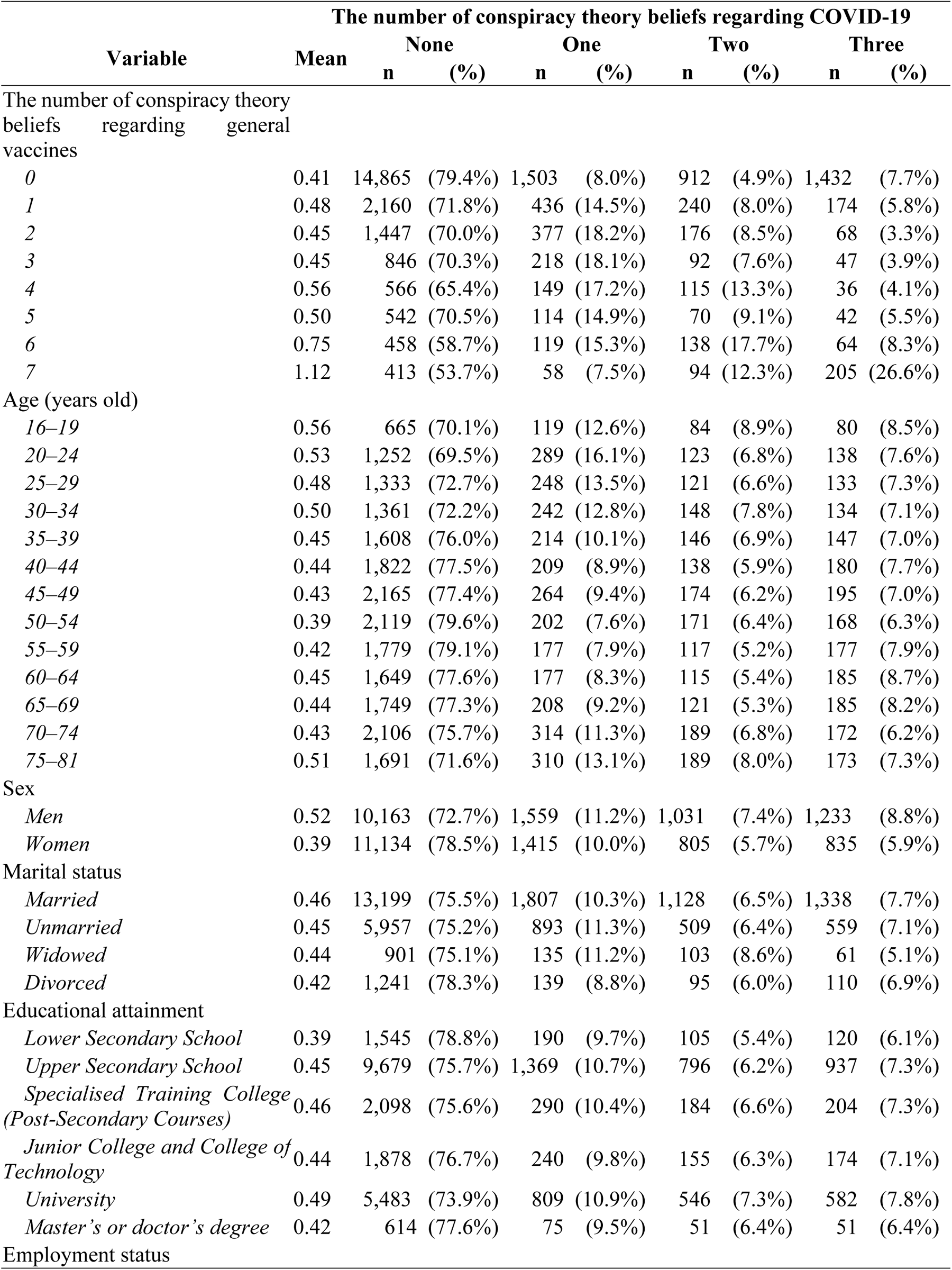
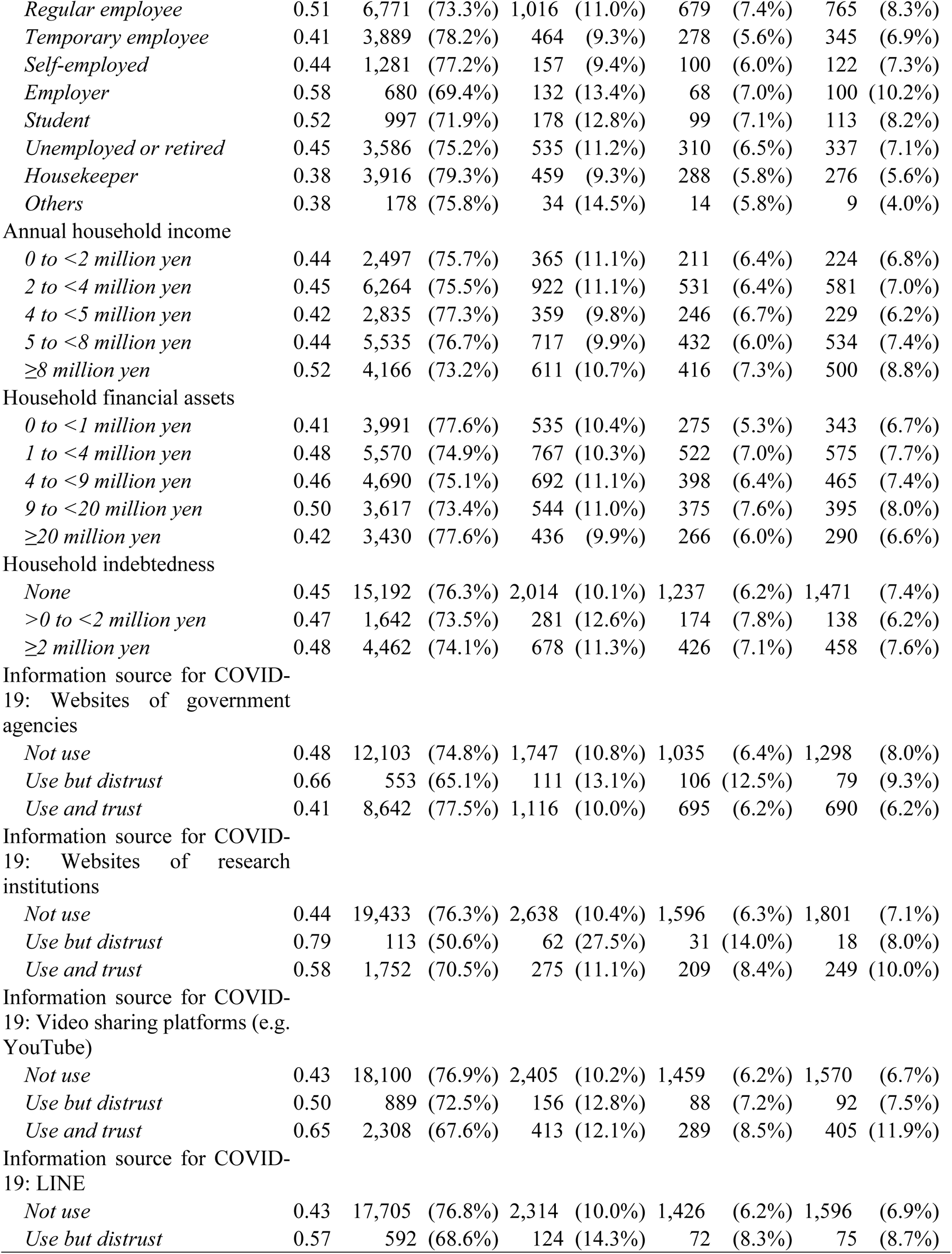
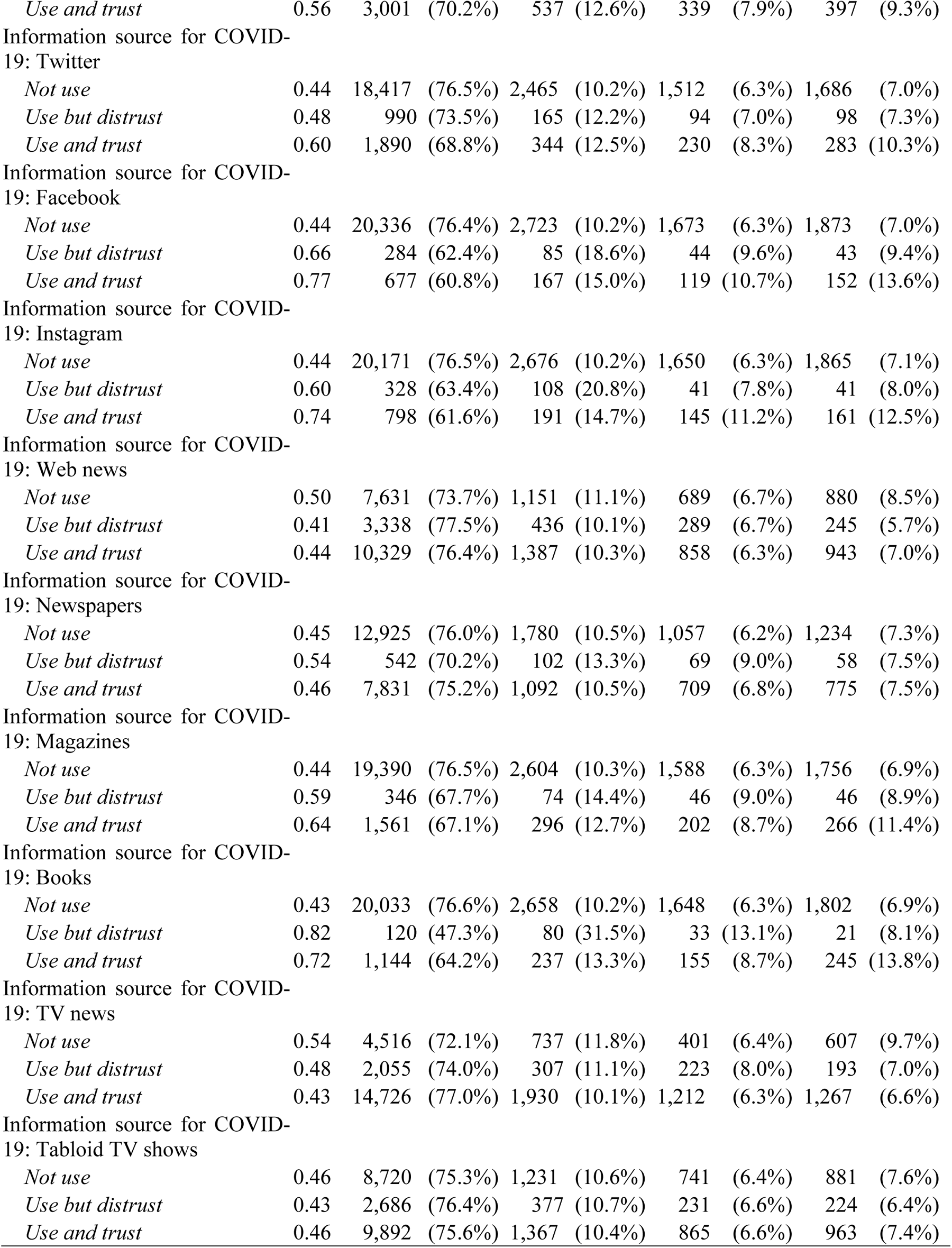
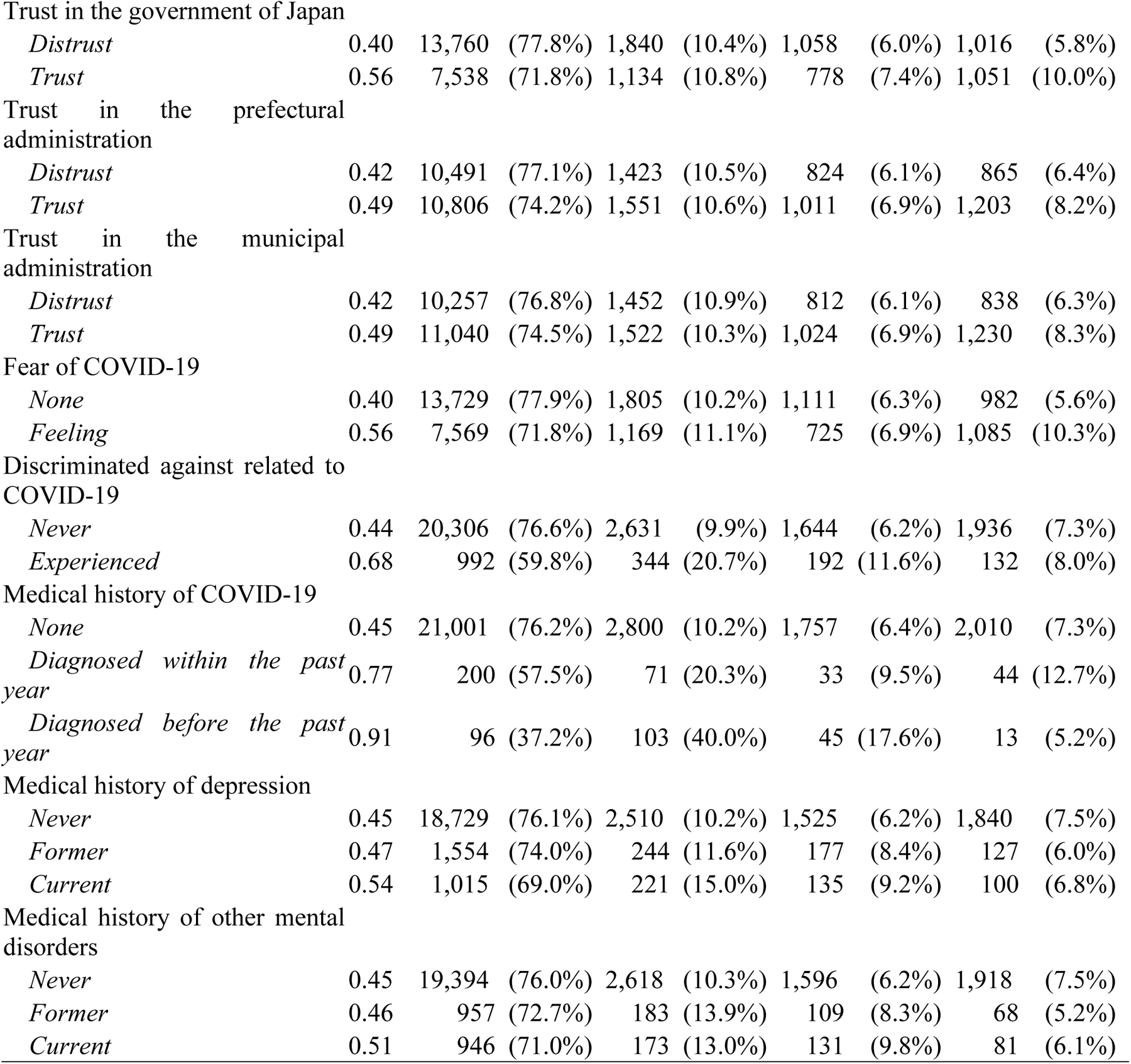
Weighted means and proportions of the number of COVID-19 conspiracy beliefs after imputation.

Table 3 shows the results of the crude and multivariable linear regression models with sampling weights after imputation. In the multivariable adjusted model, socioeconomic status was associated with harboring conspiracy theories about COVID-19. Lower secondary education and a master’s or doctor’s degree were associated with endorsing fewer conspiracy theories about COVID-19 compared with upper secondary education (lower secondary education: B -0.089, 95%CI -0.172, -0.006; master’s or doctor’s degree: B -0.066, 95%CI - 0.126, -0.006). Temporary employees (B -0.051, 95%CI -0.095, -0.007), the self-employed (B -0.102, 95%CI -0.162, -0.042), the unemployed or retired (B -0.078, 95%CI -0.131, -0.025), and housekeepers (B -0.086, 95%CI -0.135, -0.037) held fewer conspiracy theories than regular employees. In addition, higher household income and assets were also associated with endorsing more conspiracy theories (annual household income of ≥8 million yen: B 0.069, 95%CI 0.020, 0.117 vs. 4 to <5 million yen; household financial assets of 9 to <20 million yen: B 0.060, 95%CI 0.010, 0.109 vs. 0 to <1 million yen).

**Table 3.** Associations between independent variables and the number of COVID-19 conspiracy beliefs.

| Variable | Weighted crude model<br>(n=28,175) |  | Weighted multivariable adjusted model<br>(n=28,175) |  |
| --- | --- | --- | --- | --- |
|  | B | 95% CI | B | 95% CI |
| The number of conspiracy theory beliefs regarding general vaccines |  |  |  |  |
| 0 (reference) | — | — | — | — |
| 1 | 0.071 | 0.027, 0.115 | 0.059 | 0.017, 0.102 |
| 2 | 0.043 | -0.008, 0.093 | 0.045 | -0.004, 0.094 |
| 3 | 0.043 | -0.021, 0.107 | 0.040 | -0.022, 0.103 |
| 4 | 0.154 | 0.073, 0.234 | 0.178 | 0.098, 0.258 |
| 5 | 0.088 | 0.002, 0.174 | 0.128 | 0.042, 0.215 |
| 6 | 0.347 | 0.243, 0.451 | 0.401 | 0.299, 0.503 |
| 7 | 0.710 | 0.585, 0.835 | 0.718 | 0.593, 0.843 |
| Age (years old) |  |  |  |  |
| 45–49 (reference) | — | — | — | — |
| 16–19 | 0.129 | 0.012, 0.246 | 0.140 | 0.013, 0.266 |
| 20–24 | 0.098 | 0.029, 0.167 | 0.080 | -0.002, 0.163 |
| 25–29 | 0.057 | -0.011, 0.125 | 0.029 | -0.040, 0.098 |
| 30–34 | 0.071 | 0.002, 0.139 | 0.060 | -0.008, 0.128 |
| 35–39 | 0.020 | -0.045, 0.085 | 0.027 | -0.036, 0.090 |
| 40–44 | 0.009 | -0.054, 0.071 | 0.001 | -0.060, 0.061 |
| 50–54 | -0.033 | -0.095, 0.028 | -0.025 | -0.085, 0.034 |
| 55–59 | -0.009 | -0.072, 0.053 | -0.004 | -0.066, 0.058 |
| 60–64 | 0.024 | -0.040, 0.089 | 0.050 | -0.015, 0.116 |
| 65–69 | 0.016 | -0.047, 0.080 | 0.053 | -0.014, 0.121 |
| 70–74 | 0.006 | -0.054, 0.067 | 0.050 | -0.016, 0.116 |
| 75–81 | 0.083 | 0.013, 0.153 | 0.095 | 0.015, 0.174 |
| Sex |  |  |  |  |
| Men (reference) | — | — | — | — |
| Women | -0.133 | -0.162, -0.105 | -0.095 | -0.130, -0.061 |
| Marital status |  |  |  |  |
| Married (reference) | — | — | — | — |
| Unmarried | -0.009 | -0.042, 0.024 | -0.054 | -0.095, -0.013 |
| Widowed | -0.026 | -0.100, 0.049 | -0.012 | -0.089, 0.064 |
| Divorced | -0.047 | -0.107, 0.014 | -0.032 | -0.096, 0.031 |
| Educational attainment |  |  |  |  |
| Upper Secondary School (reference) | — | — | — | — |
| Lower Secondary School | -0.064 | -0.150, 0.023 | -0.089 | -0.172, -0.006 |
| Specialised Training College (Post-Secondary Courses) | 0.005 | -0.039, 0.050 | 0.018 | -0.026, 0.061 |
| Junior College and College of Technology | -0.013 | -0.056, 0.029 | 0.034 | -0.008, 0.077 |
| University | 0.040 | 0.010, 0.070 | 0.009 | -0.024, 0.042 |
| Master's or doctor's degrees | -0.035 | -0.092, 0.022 | -0.066 | -0.126, -0.006 |
| Employment status |  |  |  |  |
| Regular employee (reference) | — | — | — | — |
| Temporary employee | -0.093 | -0.134, -0.052 | -0.051 | -0.095, -0.007 |
|  | B | 95% CI | B | 95% CI |
| *Self-employed* | -0.071 | -0.132, -0.010 | -0.102 | -0.162, -0.042 |
| *Employer* | 0.075 | -0.011, 0.161 | 0.004 | -0.081, 0.089 |
| *Student* | 0.010 | -0.071, 0.091 | -0.061 | -0.155, 0.034 |
| *Unemployed or retired* | -0.052 | -0.096, -0.008 | -0.078 | -0.131, -0.025 |
| *Housekeeper* | -0.129 | -0.167, -0.091 | -0.086 | -0.135, -0.037 |
| *Others* | -0.126 | -0.257, 0.004 | -0.118 | -0.248, 0.012 |
| Annual household income |  |  |  |  |
| *4 to <5 million yen (reference)* | — | — | — | — |
| *0 to <2 million yen* | 0.023 | -0.038, 0.085 | 0.038 | -0.024, 0.101 |
| *2 to <4 million yen* | 0.030 | -0.014, 0.074 | 0.035 | -0.008, 0.078 |
| *5 to <8 million yen* | 0.022 | -0.021, 0.065 | 0.016 | -0.026, 0.058 |
| *≥8 million yen* | 0.098 | 0.050, 0.145 | 0.069 | 0.020, 0.117 |
| Household financial assets |  |  |  |  |
| *0 to <1 million yen (reference)* | — | — | — | — |
| *1 to <4 million yen* | 0.065 | 0.019, 0.111 | 0.039 | -0.006, 0.084 |
| *4 to <9 million yen* | 0.051 | 0.005, 0.096 | 0.020 | -0.026, 0.066 |
| *9 to <20 million yen* | 0.092 | 0.044, 0.139 | 0.060 | 0.010, 0.109 |
| *≥20 million yen* | 0.005 | -0.042, 0.051 | -0.016 | -0.068, 0.036 |
| Household indebtedness |  |  |  |  |
| *None (reference)* | — | — | — | — |
| *>0 to <2 million yen* | 0.020 | -0.036, 0.075 | -0.025 | -0.080, 0.030 |
| *≥2 million yen* | 0.035 | 0.002, 0.069 | -0.016 | -0.052, 0.020 |
| Information source for COVID-19: Websites of government agencies |  |  |  |  |
| *Not used (reference)* | — | — | — | — |
| *Use but distrust* | 0.184 | 0.088, 0.281 | 0.083 | -0.011, 0.178 |
| *Use and trust* | -0.066 | -0.094, -0.038 | -0.109 | -0.139, -0.079 |
| Information source for COVID-19: Websites of research institutions |  |  |  |  |
| *Not used (reference)* | — | — | — | — |
| *Use but distrust* | 0.352 | 0.192, 0.512 | 0.036 | -0.125, 0.198 |
| *Use and trust* | 0.138 | 0.083, 0.193 | 0.039 | -0.016, 0.094 |
| Information source for COVID-19: Video sharing platforms (e.g. YouTube) |  |  |  |  |
| *Not used (reference)* | — | — | — | — |
| *Use but distrust* | 0.070 | -0.002, 0.143 | -0.002 | -0.073, 0.070 |
| *Use and trust* | 0.220 | 0.170, 0.270 | 0.089 | 0.038, 0.139 |
| Information source for COVID-19: LINE |  |  |  |  |
| *Not used (reference)* | — | — | — | — |
| *Use but distrust* | 0.139 | 0.050, 0.228 | 0.064 | -0.029, 0.157 |
| *Use and trust* | 0.131 | 0.088, 0.173 | 0.053 | 0.009, 0.097 |
| Information source for COVID-19: Twitter |  |  |  |  |
| *Not used (reference)* | — | — | — | — |
| *Use but distrust* | 0.043 | -0.025, 0.112 | -0.002 | -0.069, 0.066 |
| *Use and trust* | 0.164 | 0.111, 0.217 | 0.048 | -0.005, 0.102 |
|  | B | 95% CI | B | 95% CI |
| Information source for COVID-19: Facebook |  |  |  |  |
| *Not used (reference)* | — | — | — | — |
| *Use but distrust* | 0.222 | 0.107, 0.337 | 0.042 | -0.079, 0.163 |
| *Use and trust* | 0.332 | 0.246, 0.419 | 0.089 | 0.000, 0.178 |
| Information source for COVID-19: Instagram |  |  |  |  |
| *Not used (reference)* | — | — | — | — |
| *Use but distrust* | 0.165 | 0.066, 0.264 | -0.012 | -0.120, 0.095 |
| *Use and trust* | 0.306 | 0.225, 0.387 | 0.093 | 0.008, 0.178 |
| Information source for COVID-19: Web news |  |  |  |  |
| *Not used (reference)* | — | — | — | — |
| *Use but distrust* | -0.093 | -0.135, -0.052 | -0.080 | -0.123, -0.037 |
| *Use and trust* | -0.060 | -0.092, -0.029 | -0.068 | -0.102, -0.034 |
| Information source for COVID-19: Newspapers |  |  |  |  |
| *Not used (reference)* | — | — | — | — |
| *Use but distrust* | 0.092 | 0.001, 0.183 | -0.035 | -0.128, 0.057 |
| *Use and trust* | 0.018 | -0.011, 0.047 | 0.011 | -0.023, 0.045 |
| Information source for COVID-19: Magazines |  |  |  |  |
| *Not used (reference)* | — | — | — | — |
| *Use but distrust* | 0.156 | 0.053, 0.259 | 0.004 | -0.103, 0.111 |
| *Use and trust* | 0.208 | 0.151, 0.265 | 0.051 | -0.007, 0.109 |
| Information source for COVID-19: Books |  |  |  |  |
| *Not used (reference)* | — | — | — | — |
| *Use but distrust* | 0.385 | 0.239, 0.532 | 0.137 | -0.011, 0.286 |
| *Use and trust* | 0.286 | 0.219, 0.352 | 0.136 | 0.068, 0.204 |
| Information source for COVID-19: TV news |  |  |  |  |
| *Not used (reference)* | — | — | — | — |
| *Use but distrust* | -0.057 | -0.112, -0.001 | -0.040 | -0.102, 0.022 |
| *Use and trust* | -0.110 | -0.147, -0.073 | -0.081 | -0.128, -0.035 |
| Information source for COVID-19: Tabloid TV shows |  |  |  |  |
| *Not used (reference)* | — | — | — | — |
| *Use but distrust* | -0.033 | -0.077, 0.011 | 0.005 | -0.043, 0.053 |
| *Use and trust* | -0.005 | -0.036, 0.025 | 0.051 | 0.015, 0.087 |
| Trust in the government of Japan |  |  |  |  |
| *Distrust (reference)* | — | — | — | — |
| *Trust* | 0.160 | 0.130, 0.191 | 0.175 | 0.139, 0.211 |
| Trust in the prefectural administration |  |  |  |  |
| *Distrust (reference)* | — | — | — | — |
| *Trust* | 0.076 | 0.048, 0.104 | 0.026 | -0.023, 0.074 |
| Trust in the municipal administration |  |  |  |  |
| *Distrust (reference)* | — | — | — | — |
| *Trust* | 0.072 | 0.044, 0.100 | 0.025 | -0.023, 0.074 |
| Fear of COVID-19 |  |  |  |  |
| *None (reference)* | — | — | — | — |
| *Feeling* | 0.161 | 0.131, 0.192 | 0.145 | 0.115, 0.175 |
| Discriminated against related to COVID-19 |  |  |  |  |
|  | B | 95% CI | B | 95% CI |
| *Never (reference)* | — | — | — | — |
| *Experienced* | 0.235 | 0.172, 0.297 | 0.088 | 0.025, 0.151 |
| Medical history of COVID-19 |  |  |  |  |
| *None (reference)* | — | — | — | — |
| *Diagnosed within the past year* | 0.325 | 0.144, 0.505 | 0.187 | 0.011, 0.363 |
| *Diagnosed before the past year* | 0.459 | 0.327, 0.590 | 0.169 | 0.024, 0.314 |
| Medical history of depression |  |  |  |  |
| *Never (reference)* | — | — | — | — |
| *Former* | 0.015 | -0.037, 0.067 | 0.010 | -0.042, 0.062 |
| *Current* | 0.087 | 0.016, 0.159 | 0.050 | -0.026, 0.127 |
| Medical history of other mental disorders |  |  |  |  |
| *Never (reference)* | — | — | — | — |
| *Former* | 0.006 | -0.057, 0.070 | -0.041 | -0.105, 0.024 |
| *Current* | 0.057 | -0.016, 0.129 | 0.007 | -0.071, 0.085 |

CI, confidence interval
Linear regression analysis with a robust error variance was used, applying sampling weights and imputation. A positive coefficient means holding more conspiracy theories while a negative coefficient means holding fewer conspiracy theories.
The fully adjusted model included all independent variables simultaneously.

Information sources for COVID-19 were associated with holding conspiracy theories about COVID-19. Participants who used and trusted the websites of government agencies had fewer conspiracy beliefs than those who did not use the websites of government (B -0.109, 95%CI -0.139, -0.079). On the other hand, participants who used and trusted video sharing platforms (e.g. YouTube) (B 0.089, 95%CI 0.038, 0.139), LINE (B 0.053, 95%CI 0.009, 0.097), Facebook (B 0.089, 95%CI 0.000, 0.178), and Instagram (B 0.093, 95%CI 0.008, 0.178) endorsed more conspiracy theories than those who did not use them. Participants who used and trusted, or distrusted web news held fewer conspiracy theories compared with those who did not use web news (use but distrust: B -0.080, 95%CI -0.123, -0.037; use and trust: B -0.068, 95%CI -0.102, -0.034). Participants who trusted the information from books held more conspiracy beliefs than those who did not use books (B 0.136, 95%CI 0.068, 0.204). While participants who trusted TV news held fewer conspiracy theories (B -0.081, 95%CI - 0.128, -0.035 vs. not used), those who trusted tabloid TV shows held more conspiracy theories (B 0.051, 95%CI 0.015, 0.087 vs. not used).

Trust in authorities was associated with endorsing conspiracy beliefs about COVID-19. Trust in the government of Japan was associated with holding more conspiracy theories (B 0.175, 95%CI 0.139, 0.211), compared to distrust. Trust in other authorities (the administration of the municipality and prefecture in which participants currently live) was not associated with holding conspiracy theories about COVID-19.

Feeling fear of COVID-19 was associated with holding more conspiracy beliefs about COVID-19 (B 0.145, 95%CI 0.115, 0.175 vs. none). A larger number of conspiracy theory beliefs regarding general vaccines were associated with holding more conspiracy theories about COVID-19. Participants who experienced discrimination against related to COVID-19 endorsed more conspiracy theories about COVID-19 (B 0.088, 95%CI 0.025, 0.151 vs. none). Having a medical history of COVID-19 was associated with holding more conspiracy theories (diagnosed within the past year: 0.187, 95%CI 0.011, 0.363, and diagnosed before the past year: 0.169, 95%CI 0.024, 0.314).

Results from Poisson regression models with a robust error variance were consistent with the results from linear regression models (see Appendix Table 7).

## DISCUSSION

From a nationwide survey involving 28,175 residents, we estimate that 24.4% of individuals in Japan held at least one conspiracy theory about COVID-19. Surprisingly, we found that the characteristics of respondents endorsing conspiracy beliefs were often the opposite of those reported in studies from western contexts. For example, people from higher socioeconomic backgrounds–such as higher income, higher wealth, and regular employment–were more likely to endorse conspiracy beliefs about COVID-19. Furthermore, 62.7% of respondents distrusted the government of Japan but they tended to hold fewer conspiracy theories about COVID-19.

### Interpretations of the results

In our study, individuals who trusted the government were more likely to believe in conspiracy theories. This finding is in contrast to previous research in other countries [10,14] and is therefore unique. A prior study from JACSIS suggests that individuals with high trust in the government of Japan tended to practice preventive measures more often than those with low trust [36]. Our supplemental analysis also found that trust in the government of Japan were associated with low vaccine hesitancy (shown in Appendix Table 8). Therefore, we expected that individuals with high trust in the government would also disbelieve conspiracy theories. However, our results suggest the opposite–individuals sceptical of the government might have been more rational and critical about conspiracy theories, but not necessarily about preventive measures. In addition, we found that holding more COVID-19 conspiracy beliefs was associated with LOWER vaccine hesitancy (Appendix Table 8). This suggests that individuals who believed in conspiracy theories about COVID-19 were more likely to follow COVID-19 measures and trust the government. It indicates that COVID-19 conspiracy beliefs might reflect not only a conspiratorial mindset but also fear of COVID-19. Indeed, fear of COVID-19 was significantly correlated with COVID-19 conspiracy beliefs (Table 3). However, if respondents simultaneously endorsed more general vaccine and COVID-19 conspiracy beliefs, holding more COVID-19 conspiracy beliefs was associated with vaccine hesitancy (Appendix Table 9). This suggests that the Japanese government will not succeed in increasing vaccine uptake by combating conspiracy beliefs about COVID-19 alone. A broader range of conspiracy theories needs to be considered, including conspiracy theories about vaccines.

Prior studies also suggest that individuals with high socioeconomic status are more likely to disbelieve conspiracy theories [10,14]. Indeed, our results suggest that the highest educational attainment (a master’s or doctor’s degree) was associated with holding fewer conspiracy theories about COVID-19 compared with the middle educational attainment (upper secondary school, which is the most standard educational attainment in Japan). However, individuals with the lowest educational attainment (lower secondary school) also endorsed fewer conspiracy theories than those with middle educational attainment. In addition, higher socioeconomic position–based on higher income, higher wealth, and regular employment–were associated with endorsing more conspiracy theories about COVID-19. This may reflect the nature of Japanese society. In Japanese society, it has been previously noted that conspiracy beliefs often attract people from higher socioeconomic backgrounds [37]. For example, the religious cult, Aum Shinrikyo (responsible for the 1995 Tokyo subway sarin attack) promulgated antisemitic conspiracy theories, claiming that the government of Japan was secretly manipulated by Jews and Freemasons [38]. Members of the cult who planned and participated in the sarin gas attack were drawn from the elite including those with expertise in medicine, biology, chemistry, physics, and engineering [39]. Future research must identify the mechanisms behind this unique Japanese result.

The results regarding information sources in this study, as well as those of previous research [10,14,15], indicated that individuals trusting social media (Twitter, Facebook, and Instagram) were more likely to believe in conspiracy theories while those who believed in mainstream sources such as government and traditional media news tended to endorse fewer conspiracy theories. This finding may be attributed to the proliferation of conspiracy theories on social media also in Japan [40]. However, it’s important to note that the causal relationship between social media use and conspiracy beliefs is still controversial [5,15]. Regarding TV programs in Japan, trusting TV news was associated with disbelief in conspiracy theories. Although TV news has provided biased reports on vaccines; however, in the COVID-19 case, they seem to reflect the opinions of the government, authorities, and expert groups [41]. However, tabloid TV shows featured celebrities, critics, journalists, and experts who were unfamiliar with public health [42]. Therefore, the observed difference between TV news and tabloid TV shows could be due to these features. Believing in information from books was associated with endorsing conspiracy theories about COVID-19. Misinformation, especially about healthcare, thrives on books in Japan [43–45]. Because these books were bestsellers, some people might have purchased and believed them. On the other hand, individuals who believe in conspiracy theories may have actively purchased books supporting their beliefs. The correlation between books and endorsing conspiracy theories might be attributed to the proliferation of books containing misinformation.

Younger and older people believed more conspiracy theories about COVID-19. We also observed sex differences in believing conspiracy theories. Experiencing discrimination related to COVID-19 was associated with belief in conspiracy theories. In a study of gay men in the UK, experiences of discrimination led to beliefs in conspiracy theories [12,23]. It’s possible that in this case, experiencing discrimination makes individuals more likely to believe conspiracy theories. However, it could also be that those who believe in conspiracy theories simply over-reported instances of discrimination related to COVID-19. The diagnosis of COVID-19 was also associated with conspiracy beliefs. People who believe in conspiracy theories tend to engage in higher-risk behaviours and report not following government guidelines [10–14]. Consequently, a higher proportion of COVID-19 patients could have been found to believe in conspiracy theories, suggesting an observed association.

### Strengths and limitations

The limitations of this study should be noted. First, this study was cross-sectional, which means that a temporal relationship between the independent variables and the outcome remains unclear. Second, our examination of conspiracy theories about COVID-19 relied on three questions. The estimated prevalence of conspiracy beliefs about COVID-19 may be underestimated. However, the estimated prevalence was relatively similar to results from previous systematic reviews [9,10]. This study also has strengths. Unlike many previous studies with sample sizes over 200 [14], our study included 28,175 participants. In addition, the data includes a diverse range of Japanese residents, especially those with low socioeconomic status, employing sampling weights to improve external validity. Moreover, this study examined numerous potential associated factors, enabling a comprehensive assessment of the characteristics of individuals in Japan who hold conspiracy theories about COVID-19 and the results can be applied to the general population in Japan.

### Conclusions

Conspiracy theories are a global problem and pose a public health risk. This study reported the prevalence of conspiracy beliefs about COVID-19 and associated factors in Japan. Some of the results were different, while some replicated findings from Western countries. Similar to findings from previous research, individuals who used social media as a source of information endorsed conspiracy theories. However, contrary to the results from Western countries, individuals with the lowest level of education were less likely to believe in conspiracy theories. Furthermore, higher socioeconomic Japanese–higher income, higher wealth, and regular employment–are more likely to endorse conspiracy beliefs about COVID-Those who trusted the government were more likely to hold conspiracy beliefs. Our findings might support that combating the spread of conspiracy theories requires interventions that take into account cultural contexts.

## Data Availability

Availability of data and material The data used in this study are not available in a public repository because they contain personally identifiable or potentially sensitive patient information. Based on the regulations for ethical guidelines in Japan, the Research Ethics Committee of the Osaka International Cancer Institute has imposed restrictions on the dissemination of the data collected in this study. All data enquiries should be addressed to the person responsible for data management, Dr. Takahiro Tabuchi at the following e-mail address: tabuchitak@ gmail.com

## Acknowledgements

We thank all the participants who voluntarily shared their time and experience for the JACSIS.

## DECLARATIONS

### Authors’ contributions

Study concept and design: Takahiro Tabuchi and Yukihiro Sato. Acquisition of data: Takahiro Tabuchi. Analysis and interpretation of data: All authors. Drafting of the manuscript: Yukihiro Sato. Critical revision of the manuscript for important intellectual content: All authors. Final approval of the version to be published: All authors Agreement to be accountable for all aspects of the work: All authors.

### Availability of data and material

The data used in this study are not available in a public repository because they contain personally identifiable or potentially sensitive patient information. Based on the regulations for ethical guidelines in Japan, the Research Ethics Committee of the Osaka International Cancer Institute has imposed restrictions on the dissemination of the data collected in this study. All data enquiries should be addressed to the person responsible for data management, Dr. Takahiro Tabuchi at the following e-mail address: tabuchitak@ gmail.com

